# Ivermectin for the treatment of COVID-19: A systematic review and meta-analysis of randomized controlled trials

**DOI:** 10.1101/2021.05.21.21257595

**Authors:** Yuani M. Roman, Paula Alejandra Burela, Vinay Pasupuleti, Alejandro Piscoya, Jose E. Vidal, Adrian V. Hernandez

## Abstract

**Background:** We systematically assessed benefits and harms of the use of ivermectin (IVM) in COVID-19 patients.

**Methods:** Published and preprint randomized controlled trials (RCTs) assessing IVM effects on COVID-19 adult patients were searched until March 15, 2021 in five engines. Primary outcomes were all-cause mortality, length of stay (LOS), and adverse events (AE). Secondary outcomes included viral clearance and severe AEs. We evaluated risk of bias (RoB) using the Cochrane RoB 2·0 tool. Inverse variance random effect meta-analyses were performed with quality of evidence (QoE) evaluated using GRADE methodology. Subgroup analyses by severity of disease and RoB, and sensitivity analyses by time of follow-up were conducted.

**Results:** Ten RCTs (n=1173) were included. Controls were standard of care [SOC] in five RCTs and placebo in five RCTs. RCTs sample size ranged from 24 to 398 patients, mean age from 26 to 56 years-old, and severity of COVID-19 disease was mild in 8 RCTs, moderate in one RCT, and mild and moderate in one RCT. IVM did not reduce all-cause mortality vs. controls (RR 0.37, 95%CI 0.12 to 1.13, very low QoE). IVM did not reduce LOS vs. controls (MD 0.72 days, 95%CI -0.86 to 2.29, very low QoE). AEs, severe AE and viral clearance were similar between IVM and controls (low QoE for these three outcomes). Subgroups by severity of COVID-19 or RoB were mostly consistent with main analyses; all-cause mortality in three RCTs at high RoB was reduced with IVM. Sensitivity analyses excluding RCTs with follow up <21 days showed no difference in all-cause mortality.

**Conclusions:** In comparison to SOC or placebo, IVM did not reduce all-cause mortality, length of stay or viral clearance in RCTs in COVID-19 patients with mostly mild disease. IVM did not have effect on AEs or SAEs. IVM is not a viable option to treat COVID-19 patients.

## Introduction

The coronavirus disease 2019 (COVID-19) pandemic represents a sanitary, social and economic challenge at a global level. Advances in only one year about epidemiology, prevention and clinical management on COVID-19 have been unprecedented. However, those scientific advances have also amplified deficiencies and misinformation in the scientific research environment [1]. Biological plausibility, pathophysiological considerations, *in vitro* research, observational studies, and/or clinical trials with heterogeneous quality have evaluated several repurposed drugs outside the scope of the initial approved medical use. During the pandemic, some policy-makers and regulatory institutions authorized emergency use of unproven COVID-19 treatments; the use of some of these treatments has been heavily politicized in some regions of the world [2,3].

Ivermectin (IVM) is a semisynthetic, anthelmintic agent for oral administration. IVM is derived from the avermectins, which are isolated from the fermentation products of *Streptomyces avermitilis*. IVM and its analogs selectively open inhibitory glutamate-gated chloride ion channels in the membranes of pharyngeal muscles, motor nerves, female reproductive tracts, and the excretory/secretory pores of nematodes. In addition, IVM prevents the filarial ability to release substances that inhibit the host immune response[4]. In tissue cultures, at concentrations higher than anthelmintic concentrations, IVM showed antiviral (e.g., dengue), antiparasitic (e.g., malaria), and anticancer effects (e.g., epithelial ovarian cancer). However, these *in vitro* results have not been clinically demonstrated [4].

In March 2020, researchers from Australia showed IVM to be active against SARS-CoV-2 in cell cultures by drastically reducing viral RNA at 48 hours [5]. Concentrations tested in these *in vitro* assays are equivalent to more than 50-fold the normal C_max_ achieved with a standard single dose of IVM 200 μg/kg, raising concerns about the effective dose of IVM for treating or preventing SARS-CoV-2 infection in humans and its tolerability [6]. However, additional theoretical considerations, experimental and observational evidence, misinformation, self-medication and the wide availability of IVM, led to its use for the treatment of COVID-19, particularly in low- and middle-income countries, assuming *a priori* efficacy and safety.

Ivermectin is currently approved by the Food and Drug Administration (FDA) to treat people with intestinal strongyloidiasis and onchocerciasis. The European Medicines Agency [7] and FDA [8] have not approved IVM for the treatment of COVID-19. World Health Organization (WHO)[9] and Infectious Diseases Society of America (IDSA)[10] guidelines, do not recommend IVM for treatment of COVID-19 outside randomized controlled trials (RCTs).

High-quality RCTs and systematic reviews are necessary to evaluate efficacy of IVM in COVID-19.Threesystematic reviews on the effect of IVM on clinical outcomes were published [11-13]. Padhy et al. only included three small observational studies [11].

Siemieniuk et al. conducted a living systematic review of all treatments for COVID-19, but details were scarce and quality of evidence was very low [12]. Finally, Kow et al. evaluated six RCTs, five of them from Asia and none from Latin America [13]. In addition to published studies, other systematic reviews or narrative reviews of the effects of IVM have been only disseminated as pre-prints [14-16] or only presented on websites [17-19].

We conducted a systematic review and meta-analysis to evaluate treatment effects of IVM on clinical outcomes and adverse events in people with COVID-19.

## Methods

### Sources and Searches

Two investigators (V.P., and A.V.H.) developed the search strategy, which was approved by the other investigators. We searched the following databases until March 22, 2021: PubMed-MEDLINE, EMBASE-OVID, Scopus, Web of Science, the Cochrane Library, medRxiv.org (www.medrxiv.org), Preprints (www.preprints.org), and Social Science Research Network (www.ssrn.com). The PubMed search strategy is shown in the **Supplement**.

### Selection of studies

We included RCTs in any language reporting benefit or harm outcomes of IVM as treatment in COVID-19 patients, both non-hospitalized and hospitalized, irrespective of COVID-19 severity. We excluded studies assessing prophylaxis for COVID-19 infection. Controls were standard of care (SOC) or placebo. Two investigators (YMR, AB) independently screened each record title and abstract for potential inclusion, and then assessed full texts of selected abstracts. Discrepancies were resolved through discussion or by a third investigator (AVH).

### Outcomes

Primary outcomes were all-cause mortality, length of hospital stay, and adverse events (AE). Secondary outcomes were SARS-CoV-2 clearance on respiratory samples, clinical improvement, need for mechanical ventilation, and severe adverse events (SAE). AEs and SAEs were extracted as defined by authors.

### Data Extraction

Two investigators (YMR and AB) independently extracted the following variables from studies: country(ies), sample size, dose and duration of IVM treatment, type of control group (SOC vs. placebo), COVID-19 severity, percentage of positive reverse transcription polymerase chain reaction (RT-PCR) for severe acute respiratory syndrome coronavirus 2 (SARS-CoV-2), study setting (hospitalized vs. non-hospitalized), mean age, proportion of female participants, co-morbidities such as hypertension, diabetes mellitus and cardiovascular disease, evaluated outcomes, and time of follow up. COVID-19 disease severity was defined as mild, moderate or severe according to the WHO classification [19]. Discrepancies were resolved through discussion or by two other investigators (AP, AVH).

### Risk of bias assessment

Two investigators (YMR and AB) independently assessed RoB by using the Cochrane Risk of Bias 2.0 tool for RCTs [20]; disagreements were resolved by discussion with a third investigator (AP). The RoB 2.0 tool evaluates five domains of bias: randomization process, deviations from intended interventions, missing outcome data, measurement of the outcome, and selection of the reported results. RoB per each of the five domains and overall were described as low, some concerns and high.

### Statistical analyses

We reported our systematic review according to 2009 PRISMA guidelines [22]. Inverse variance random effect meta-analyses were performed to evaluate effect of IVM vs. control on outcomes. Effects of meta-analyses were reported as relative risks (RR) for dichotomous outcomes and as mean difference (MD) for the continuous outcome length of stay. The between study variance tau^2^ was calculated with the Paule-Mandel method [23] and the 95% confidence intervals (CIs) of effects were adjusted with the Hartung-Knapp method [24].We adjusted for zero events in one or two RCT arms using the continuity correction method [25]. Heterogeneity of effects among studies was quantified with the I^2^ statistic (I^2^>60% means high heterogeneity). We pre-specified subgroup analyses by severity of COVID-19 disease and RoB; the p for interaction test <0.1 indicated effect modification by subgroup. Sensitivity analyses by excluding RCTs with shorter follow-up (i.e.<21 days) were planned for the primary outcomes. The *meta* package of R 3.5.1 (www.r-project.org) was used for meta-analyses. The quality of evidence was evaluated using the GRADE methodology, which covers five aspects: risk of bias, inconsistency, indirectness, imprecision, and publication bias [26]. Quality of evidence (QoE) was evaluated per outcome, and described in summary of findings (SoF) tables; GRADEpro GDT was used to create SoF tables [27].

## Results

### Selection of studies

Our search yielded 256 citations with an additional nine citations identified in pre-print web pages; 253 records were excluded. After our assessment of 12 full-texts, we identified 10 RCTs [28-37] (n=1,173) (**Figure 1**). Two full-texts were excluded as there was no control group in one study, and an outcome of no interest (duration of fever) was the only one reported in another study.

**Figure 1.**
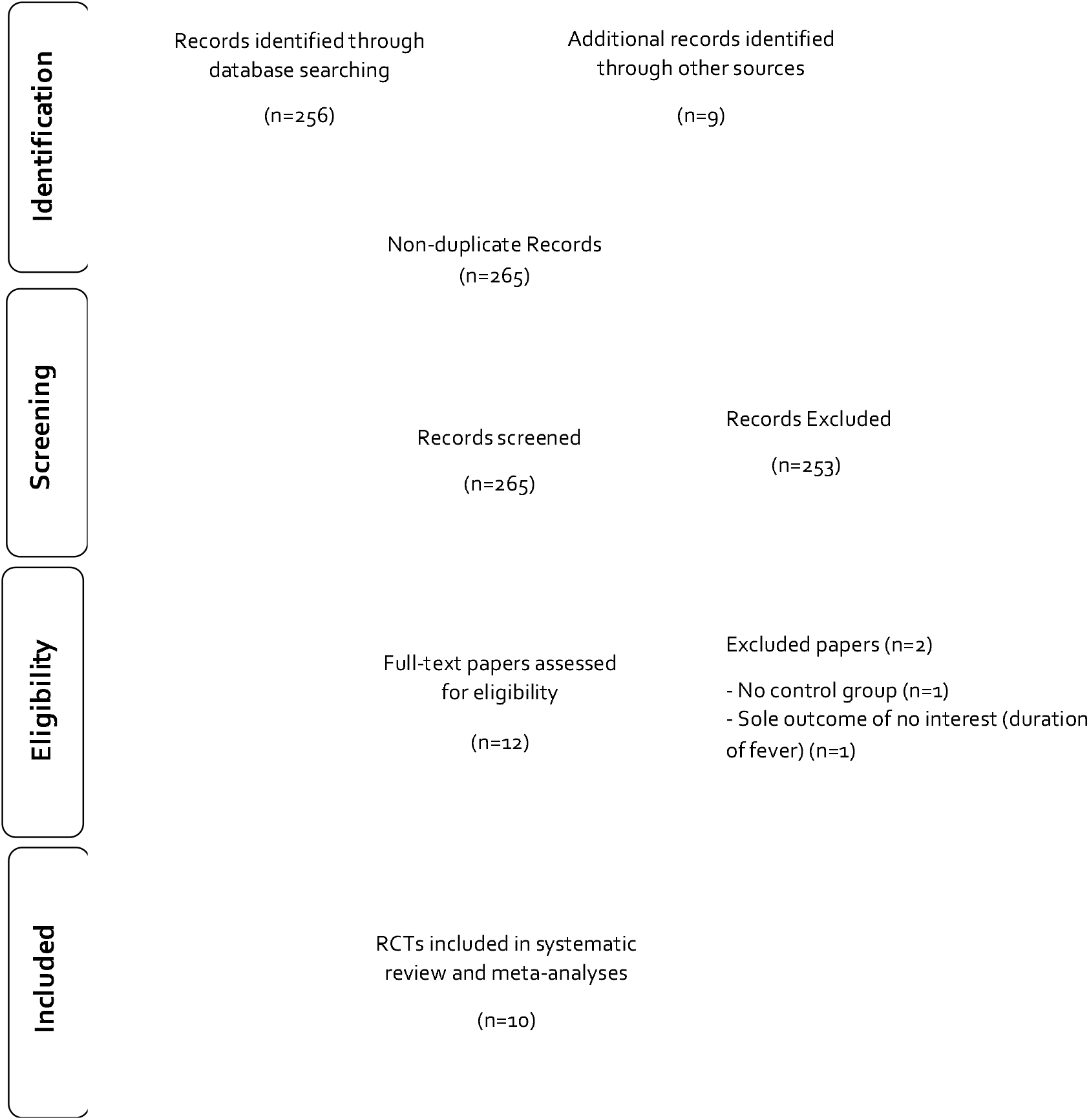
PRISMA flowchart diagram

### Characteristics of RCTs

One RCT was conducted in Spain [34] and the other nine RCTs in low- and middle-income countries. Sample sizes of RCTs ranged from 24[34] to 398 [36] patients. IVM doses were heterogeneous in terms of doses and duration (one to five days). Controls were SOC in five RCTs [28-31, 35] and placebo in five RCTs [32-34, 36, 37].Most of RCTs were conducted in mild COVID-19 patients: mild in all or the majority of patients in eight RCTs [28, 29, 31, 32, 34-37], moderate in one RCT [33], and mild and moderate in one RCT [30] (**Table 1**).

**Table 1.** Study characteristics of included randomized controlled trials.

| Study, year, reference | Country, sample size | IVM dose and duration | Control group | COVID-19 severity according to WHO classification<sup>19</sup> | Positive SARS-CoV-2 RT-PCR (%) | Hospitalized (%) | Mean (SD) or Median (IQR) Age | Female (%) | CVD or CHD (%) | DM (%) | HTN (%) | Evaluated outcomes | Follow-up time (days) |
| --- | --- | --- | --- | --- | --- | --- | --- | --- | --- | --- | --- | --- | --- |
| Chachar 2020<sup>28</sup> | Pakistan, 50 | 12mg; 12 mg at 12 h, and 12mg at 24 h. | SOC | Mild (100%) | 100 | 0 | 42 (16) | 38 | 8 | 40 | 26 | Asymptomatic at day 7 | 7 |
| Krolewiecki 2020<sup>29</sup> | Argentina, 45 | 0.6 mg/kgqd for 5 days. | SOC | Mild (87%), moderate (13%) | 100 | 100 | 41 (12) | 44 | NR | 16 | 13 | Viral load at day 5, IVM plasma level | 30 |
| Niaee 2020<sup>30</sup> | Iran, 180 | Four doses: from 200 µg/kg single dose to 800 µg/kg across 5 days. | SOC | Mild and moderate (unclear distribution) | 71 | 100 | 56 (45-67) | 50 | NR | NR | NR | All-cause mortality, time until remission of symptoms, hospital LOS | 5 |
| Podder 2020<sup>31</sup> | Bangladesh, 62 | 200 µg/kg single dose. | SOC | Mild (81%), moderate (19%) | 100 | NR | 39 (12) | 29 | NR | NR | NR | Time to full recovery, viral clearance | 10 |
| Ahmed 2021<sup>32</sup> | Bangladesh, 48 | 12 mg qd for 5 days. | Placebo | Mild (100%) | 100 | 100 | 42 (NR) | 54 | 0 | 0 | 0 | Remission of symptoms, hospital LOS, SAE, oxygen requirement, time to viral clearance | 14 |
| Beltran 2021<sup>33</sup> | Mexico, 73 | 12 mg if <80 kg; 18 mg if >80 kg, single dose | Placebo | Moderate (74% with PaO<sub>2</sub>/FiO<sub>2</sub> ratio 100 to 300) | 100 | 100 | 53 (17) | 38 | NR | 34 | 32 | All-cause mortality, clinical recovery, hospital LOS, AE, respiratory deterioration | 28 |
| Chaccour 2021<sup>34</sup> | Spain, 24 | 400 µg/kg single dose. | Placebo | Mild (100%) | 100 | 0 | 26 (19-36) | 50 | 0 | 0 | 0 | All-cause mortality, AE, PCR at day 7 | 28 |
| Karamat 2021<sup>35</sup> | Pakistan, 86 | 12 mg single dose. | SOC | Mild (most of them, unclear %) | 100 | 100 | 39 (42) | 15 | 5.8 | 12 | 14 | Time to viral clearance, AE | 28 |
| Lopez-Medina 2021<sup>36</sup> | Colombia, 398 | 300 µg/kgqd for 5 days. | Placebo | Mild (100%) | 100 | 1 | 37 (29-48) | 78 | 1.7 | 6 | 13 | All-cause mortality, time to complete resolution, AE, SAE, escalation of care | 21 |
| Ravikirti 2021<sup>37</sup> | India, 115 | 12 mgqd for 2 days. | Placebo | Mild (79%), moderate (21%) | Positive RT-PCR or RAT | 100 | 53 (15) | 28 | 11 | 36 | 35 | All-cause mortality, admission to ICU, requirement of MV, viral clearance at day 6 | 10 |

SD: Standard deviation; IQR: Interquartile range; SOC: Standard of care; qd: once a day; NR: Not reported; CVD: Cardiovascular disease; CHD: Coronary heart disease; DM: Diabetes mellitus; HTN: Hypertension; SARS-CoV-2: Severe acute respiratory syndrome coronavirus 2; RT-PCR: Reverse transcription polymerase chain reaction; RAT: Rapid antigen test; PaO<sub>2</sub>/FiO<sub>2</sub>: ratio of arterial oxygen partial pressure (PaO<sub>2</sub> in mmHg) to fractional inspired oxygen (FiO<sub>2</sub> expressed as a fraction, not a percentage); WHO: World Health Organization; AE: adverse events; SAE: severe adverse events; LOS: length of stay; MV: mechanical ventilation; ICU: Intensive care unit.

All patients had positive RT-PCR for SARS-CoV-2 at baseline, except in 2 RCTs: Niaee et al. [30] had 71% of positivity, and Ravikirti et al. [37] had positive RT-PCR or rapid antigen test for SARS-CoV-2. The RCTs by Chachar et al. [28], Chaccour et al. [34], and López-Medina et al. [36] were conducted in non-hospitalized patients. Mean/median age ranged from 26 to 56 years-old, the percentage of female patients ranged from 15% [35] to 78% [36], and most of patients did not have hypertension, diabetes or cardiovascular disease. Evaluated outcomes were also heterogeneous across RCTs, and time of follow up ranged from five [29) to 30 days [29].

### Risk of bias assessment of included RCTs

Eight RCTs were at high RoB [28-32, 35-37] (**Figure S1**). Podder et al. [31] had high RoB in the randomization process, deviations from intended interventions, missing outcome data, and measurement of the outcome; Karamat et al. [35] had high RoB in deviations from intended interventions and missing outcome data; Niaee et al. [30] had high RoB in the measurement of the outcome, and selection of the reported results; Krolewiecki et al. [29] and Ravikirti et al. [37] had high RoB in missing outcome data; Lopez-Medina et al. [36] had high RoB in deviations from the intended interventions; Ahmed et al. [32] had high RoB in selection of the reported results; and Chachar et al. [28] had high RoB in measurement of the outcome. Beltran et al. [33] had some concerns of bias in the randomization process.

### Meta-analyses

IVM did not have effect on all-cause mortality vs. controls in five RCTs (RR 0.37, 95%CI 0.12 to 1.13, I^2^=16%, very low QoE) (**Figure 2, Table 2**), on length of stay vs. controls in three RCTs (MD 0.72 days, 95%CI -0.86 to 2.29, I^2^=0%, very low QoE) (**Figure 3, Table 2**), and on adverse events vs. controls in three RCTs (RR 0.95, 95%CI 0.85 to 1.07, I^2^=0%, low QoE) (**Figure 4, Table 2**).

**Table 2.**
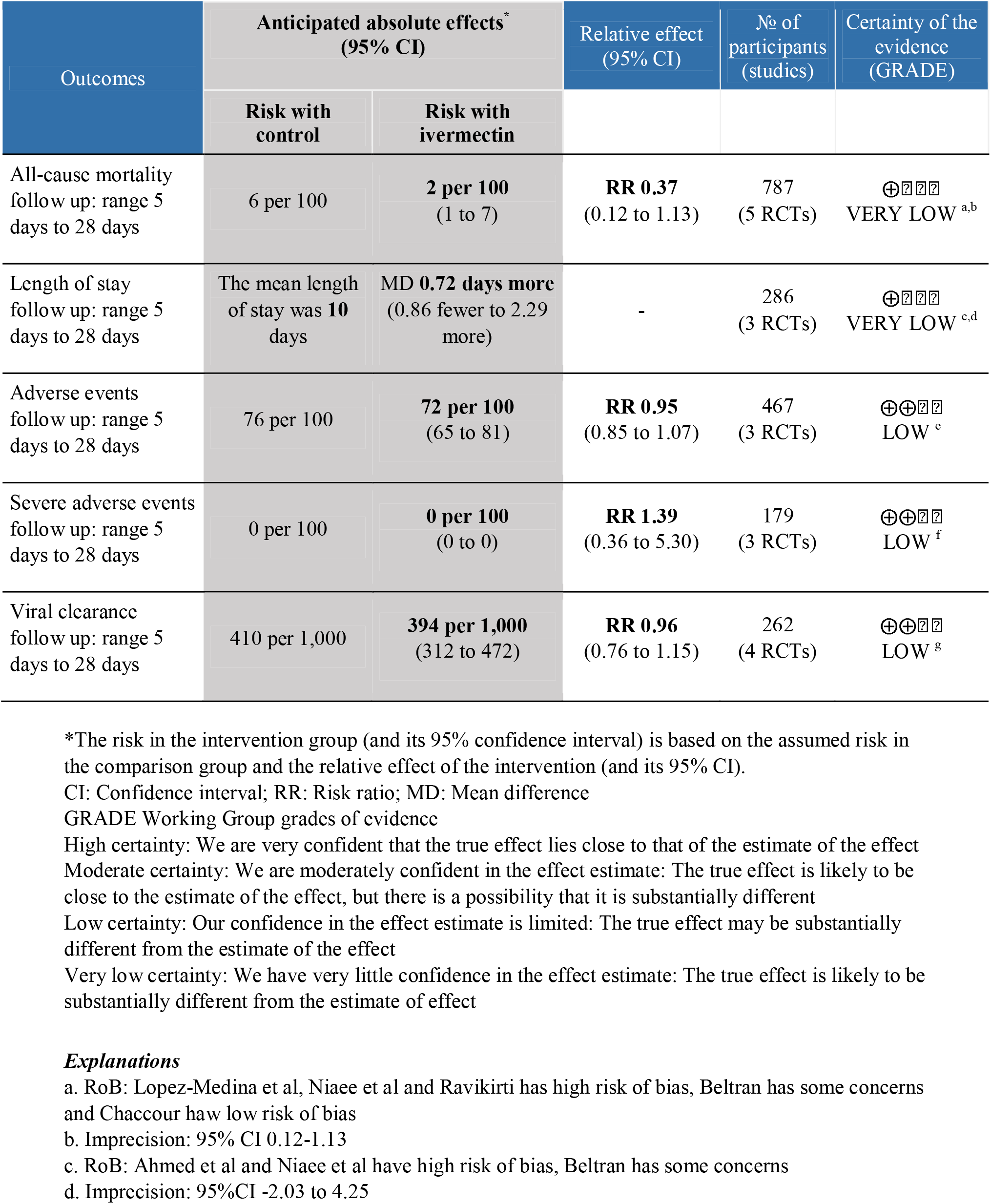
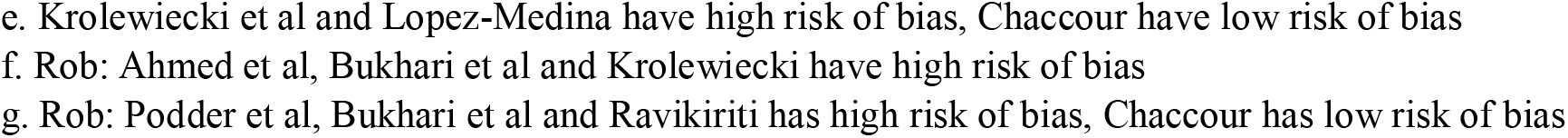
Summary of findings table of the effect of ivermectin compared to standard of care or placebo for COVID-19 patients

**Figure 2.**
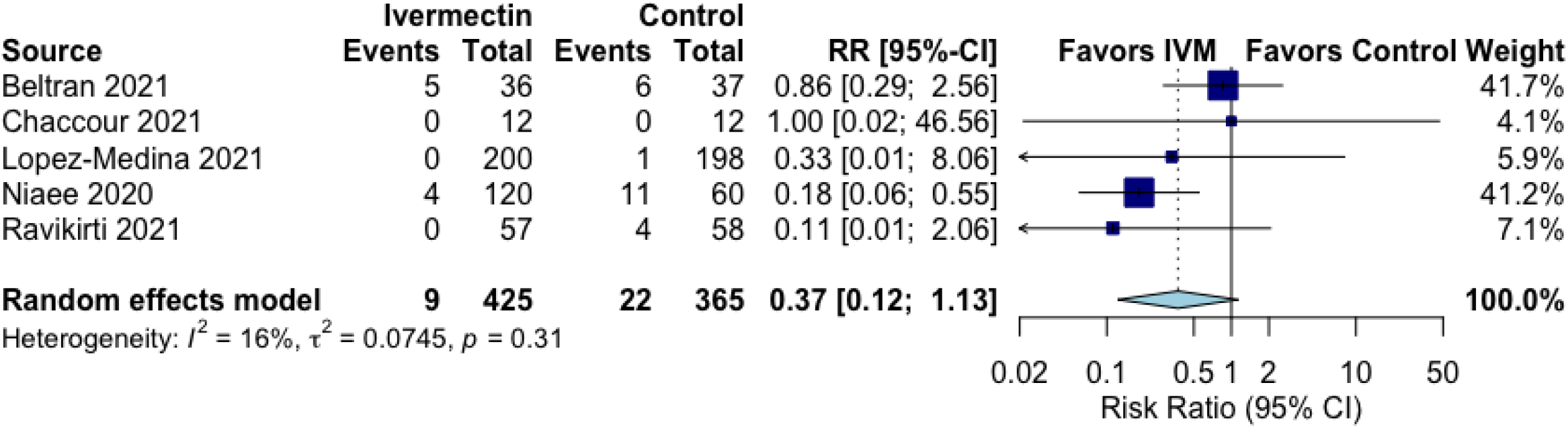
Effect of ivermectin on all-cause mortality in randomized controlled trials of COVID-19 patients.

**Figure 3.**
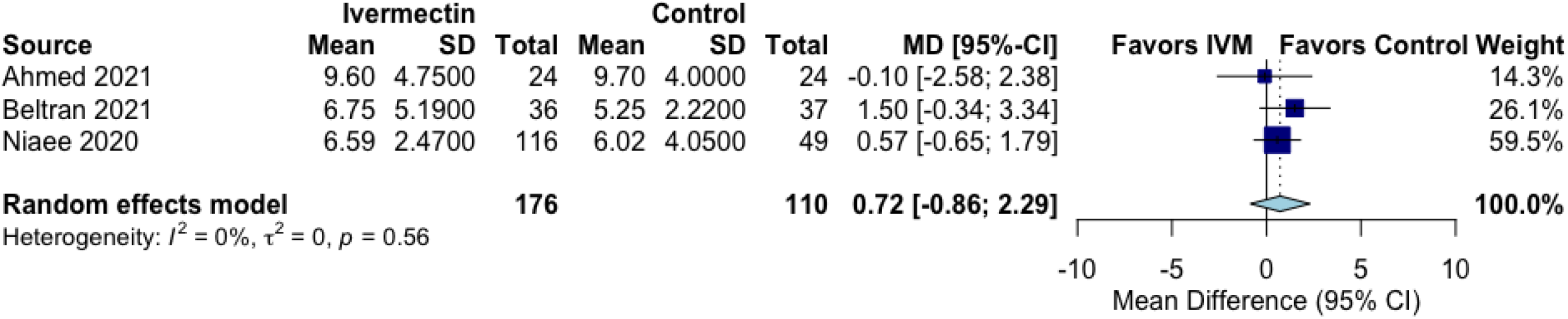
Effect of ivermectin on length of stay in randomized controlled trials of COVID-19 patients.

**Figure 4.**
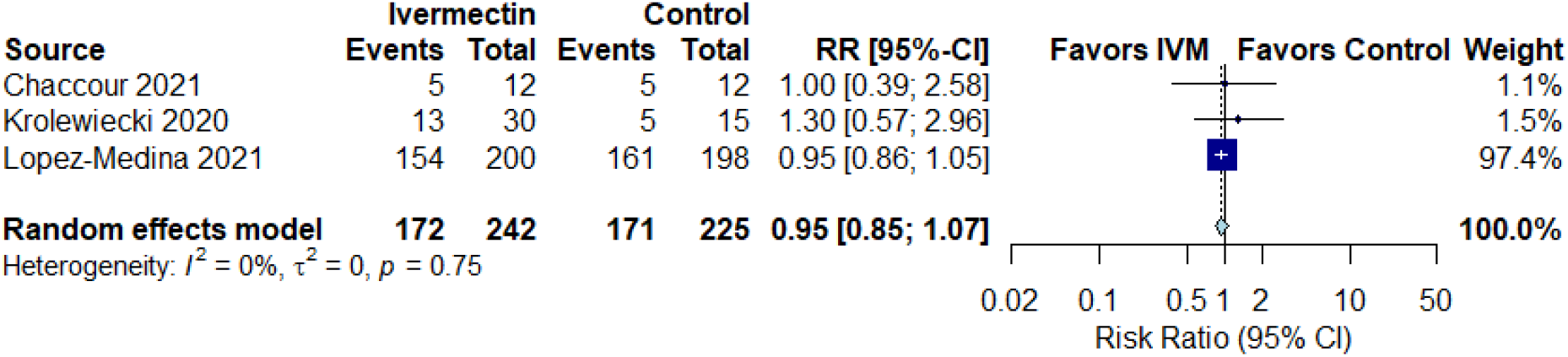
Effect of ivermectin on adverse events in randomized controlled trials of COVID-19 patients.

There was no effect of IVM on severe adverse events in comparison to the controls in three RCTs (RR 1.39, 95%CI 0.36 to 5.30, I^2^=0%, low QoE) (**Figure 5, Table 2**) and on viral clearance in comparison to the controls in four RCTs (RR 0.96, 95%CI 0.79 to 1.16, I^2^=0%, low QoE) (**Figure 6, Table 2**).

**Figure 5.**
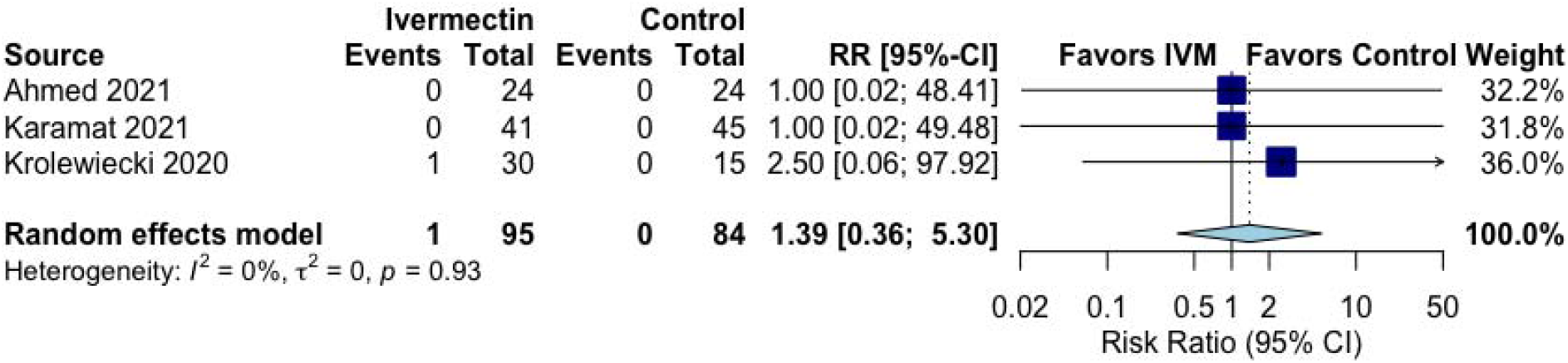
Effect of ivermectin on severe adverse events in randomized controlled trials of COVID-19 patients.

**Figure 6.**
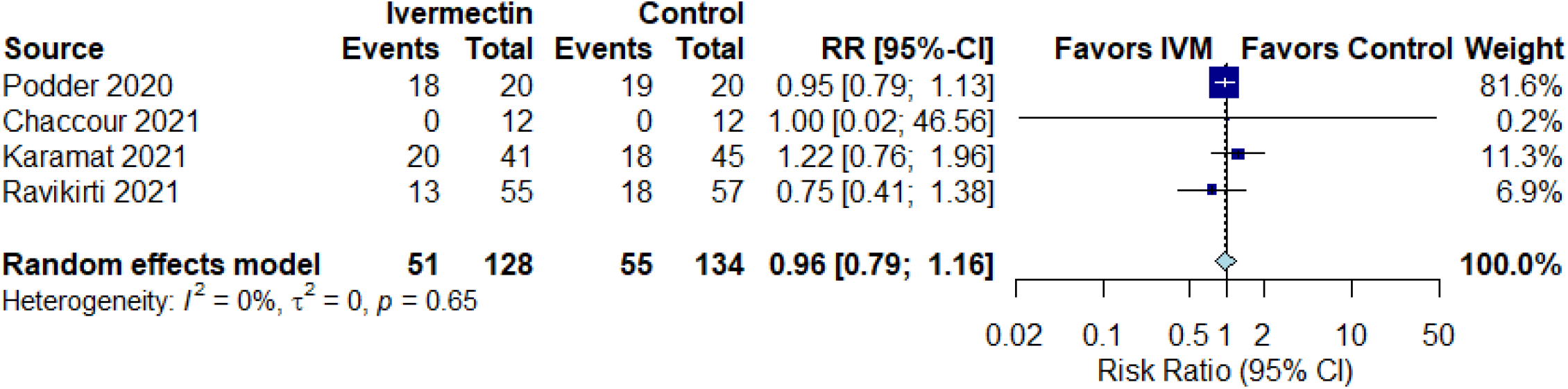
Effect of ivermectin on viral clearance in randomized controlled trials of COVID-19 patients.

### Subgroup and sensitivity analyses

Subgroup analyses by severity of COVID-19 disease or RoB were consistent with main analyses (**Figures S2.1 to S2.5**), except the subgroup by RoB of all-cause mortality; three studies [30, 36, 37] at high risk of bias showed a significant reduction in all-cause mortality (RR 0.18, 95%CI 0.07-0.49; RoB p for interaction=0.1). Sensitivity analyses excluding studies with follow-up <21 days showed similar effects as primary analyses for all-cause mortality and length of stay (**Figures S3.1 and S3.2**). Statistical heterogeneity of effects for all-cause mortality was 0% in sensitivity analysis.

## Discussion

This systematic review provides a comprehensive overview of the current evidence for IVM effects on COVID-19 patients. In comparison to SOC or placebo, IVM did not reduce the risk of primary outcomes (all-cause mortality, length of hospital stay, and adverse events) or secondary outcomes (SARS-CoV-2 clearance in respiratory samples, and severe adverse events) in RCTs of patients with mostly mild COVID-19 disease. The quality of evidence was low or very low for all outcomes. Subgroup analyses by severity of COVID-19 disease or risk of bias were mostly consistent with main analyses, except a significant effect on all-cause mortality in three RCTs at high risk of bias. Sensitivity analyses excluding RCTs with follow up <21 days showed no difference in all-cause mortality but diminished heterogeneity.

The findings of this study should be discussed in the context of prior published reports: two conventional systematic review and meta-analyses and two living systematic review and meta-analyses [9, 11-13] (**Table S1**). Padhy et al. published the first systematic review and meta-analysis about IVM in COVID-19 patients and their primary outcome was all-cause mortality [11]. This study included only 4 observational studies because none RCT had been reported and included 629 participants. IVM showed reduction of all-cause mortality (OR 0.53, 95%CI 0.09-0.36). However, the authors claim caution as the quality of evidence was very low [11]. Kow et al. published a systematic review and meta-analysis of IVM effects in COVID-19 patients on all-cause mortality [13]. This study included only 6 RCTs and 1255 participants. IVM showed reduction of all-cause mortality (OR 0.21, 95%CI 0.11-0.42).The authors showed high risk of bias in the most of the RCTs, described their findings as a preliminary positive effect, and suggested that IVM should preferably be administered under clinical trial settings until evaluated by more conclusive large-scale RCTs [13].

The WHO published a living systematic review and network meta-analysis (NMA) about IVM in COVID-19 patients with all-cause mortality as primary outcome [9]. Sixteen RCTs were evaluated, but only five directly compared IVM with SOC and reported mortality (n=915); IVM reduced all-cause mortality (OR 0.19, 95%CI 0.09-0.36). However, QoE was very low for mortality and the panel concluded that the effect of IVM on mortality was uncertain. Other outcomes (i.e. mechanical ventilation, hospital admission, and duration of hospitalization) also had very low QoE. WHO only recommended using IVM in clinical trials [9]. Siemieniuk et al. published a living systematic review and NMA about IVM in COVID-19 patients with mortality as primary outcome and other ten outcomes including hospitalization, and time to viral clearance [12]. Seven RCTs contributed to mortality assessment (n=751). IVM showed reduction of mortality (Risk difference per 1000 vs. SOC: -103, 95%CI -117 to -78), but the QoE was very low. For other critical outcomes the QoE was low; subgroup analyses did not show differential IVM effects. This study concluded that effects of IVM were highly uncertain and there was no definitive evidence of important benefits and harms for any outcomes [12]. Taken together, the results of these four studies suggested that IVM should not be used in COVID-19 patients. Living systematic reviews allow authors to update the evidence regularly, which is particularly important in a pandemic scenario [38].

In addition to fully published systematic reviews, we found three pre-prints of systematic reviews [14-16] (**Table S2**). Castañeda-Sabogal et al. evaluated 12 studies (six RCTs, five retrospective cohorts, and one case series, n=7412 overall), without description of COVID-19 severity. IVM non-significantly reduced mortality (RR 0.70, 95%CI 0.31-2.28) and non-significantly increased recovery (RR 1.37, 95%CI 0.61 to 3.07). Authors concluded that there was insufficient certainty and low QoE to recommend the use of IVM to treat COVID-19 patients [14]. Hill et al. evaluated 18 RCTs and 2282 participants with mostly mild to moderate severity. In six RCTs (four pre-prints and two trial registry web records) with 1255 participants, IVM reduced all-cause mortality (RR 0.25, 95%CI 0.12 to 0.52) but had a non-significant increase of recovery (RR 1.37, 95%CI 0.61-3.07). The quality was classified as limited in four RCTs, fair in one RCT, and good in one RCT. These authors concluded that IVM should be evaluated in well-designed, large RCTs [15]. Finally, Bryant et al. evaluated 19 RCTs (n=2003). In thirteen RCTs (three published RCTs, nine pre-prints, and one trial registry web registry) with 1892 participants with mostly mild to moderate severity, IVM reduced mortality (aRR 0.32, 95%CI 0.14 to 0.72) [16]; QoE was of low to moderate. Authors recommended the use of IVM in COVID-19, in particular in early disease without supporting data. The last two studies [15,16] used very flexible research strategies and included 0 and 13% of peer-reviewed studies, respectively. In consequence, they were subject to selection bias that explain, at least in part, the effect of IVM on mortality.

Several websites published systematic reviews and meta-analyses about IVM in COVID-19 patients with unclear or absent methodology and reporting guidelines [17-19] (**Table S2**). These websites did not include protocol registration and have relevant omissions such as inclusion criteria [19], databases searched [18, 19], quality assessment of the included studies [17,19], methods of meta-analysis [19], and definition of heterogeneity [17,19].

Arbitrarily broad inclusion criteria (i.e. studies directly submitted to the websites, more pre-prints than peer-review studies) led to a high number of RCTs and participants. For example, a “real time meta-analysis” ivmmeta.com included 46 studies, 24 of them RCTs, and 15,480 participants [17]. Coincidentally, these three studies showed beneficial outcome effects with IVM [17-19]. In a context of misinformation infodemic, the dissemination of these results caused confusion for patients, clinicians (in particular those without training in critical reading of scientific literature), and decision-makers, which, additionally, may manipulate the information with political interests [39].

The irrational use of IVM to treat COVID-19 patients has demonstrated several limitations in management strategies: absence of transparency by some political leaders or media in order to avoid the use of drugs without evidence of efficacy and concerns about safety; lack of decisive leadership to implementing therapeutic science-based guidelines; and misuse of both effective communication and science of communication [40-42]. Similar issues were previously experienced with hydroxychloroquine and this situation likely will be repeated in the future with other repurposed drugs. To avoid it, there is an urgent need to establish collaborative efforts among scientists, practitioners, communicators, and policy-makers. A large, well-designed and -reported RCT provides the most reliable information of efficacy in the specific target population from which the sample was drawn. Similarly, a well-designed and reported meta-analysis can provide valuable and confirmatory information [43, 44]. This condition is relevant in a pandemic where timely evaluations are needed.

Our study has several strengths. First, we performed a recent and comprehensive systematic search in five engines and unpublished studies and we did not restrict by language. Second, we only evaluated RCTs; several previous studies included all types of designs and their findings may have been biased and confounded. Third, we evaluated outcomes with information from at least two RCTs, including all-cause mortality, length of stay, viral clearance, adverse events and severe adverse events. No data was available for clinical improvement and need for mechanical ventilation. Fourth, we described the severity of COVID-19 disease per RCT carefully, using the WHO classification [19]; our findings do not support the use of IVM in mild disease. Fifth, we performed subgroup analyses by RoB and severity of disease, which were mostly similar to main analyses; however, we found that three RCTs at high risk of bias [30, 36, 37] had a significant reduction of all-case mortality. Sixth, we also performed sensitivity analysis by excluding studies with short follow up times; effects were similar. Finally, we evaluated the quality of evidence using GRADE methodology.

Our study also has some limitations. First, quality of evidence was low or very low for all outcomes. However, our systematic review and meta-analysis evaluated the best current available evidence and all IVM effects were negative. Second, we included only ten RCTs, five of them using a placebo as control group, and studies included a relative low number of participants. However, included RCTs are the available studies until March 15, 2021. Third, all selected RCTs evaluated patients with mild or mild to moderate COVID-19. However, the supposed benefit of IVM has been positioned precisely for mild disease, but we did not find differential IVM effects between these two severity categories. Finally, four of the RCTs had a follow-up time of only 5-10 days; analyses of primary outcomes excluding these short follow up studies showed similar IVM effects.

In conclusion, in comparison to SOC or placebo, IVM did not reduce all-cause mortality, length of stay, respiratory viral clearance, adverse events and serious adverse events in RCTs of patients with mild to moderate COVID-19. We did not find data about IVM effects on clinical improvement and need for mechanical ventilation. Additional ongoing RCTs should be completed in order to update our analyses. In the meanwhile, IVM is not a viable option to treat COVID-19 patients and only should be used within clinical trials context. In the scenario of the current COVID-19 pandemic and misinformation infodemic, our systematic review and meta-analysis provides valuable information for clinicians, researchers, and policy-makers.

## Data Availability

All data we used for this manuscript are available in text, tables, figures and supplementary file.

## Notes

**No conflicts of interest for all authors**

### Competing Interest Statement

The authors have declared no competing interest.

### Funding Statement

No external funding was received for this manuscript.

### Author Declarations

No ethics committee approval was necessary as this was a systematic review.

## References

1. Anonymous. Science during COVID-19: where do we go from here. Lancet.2021; 396(10267): 1941. DOI: 10.1016/S0140-6736(20)32709-4.

2. Saag MS. Misguided Use of Hydroxychloroquine for COVID-19. The infusion of Politics Into Science. JAMA. 2020; 324(21): 2161–2. DOI: doi: 10.1001/jama.2020.22389.

3. Barberia LG, Gómez EJ. Political and institutional perils of Brazil’s COVID-19 crisis. Lancet. 2020; 396(10248): 367–368. DOI: 10.1016/S0140-6736(20)31681-0.

4. Martin RJ, Robertson AP, Choudhary S.Ivermectin: An Anthelmintic, an Insecticide, and Much More. TrendsParasitol. 2021; 37(1): 48–64. DOI: 10.1016/j.pt.2020.10.005.

5. Chaccour C, Hammann F, Ramón-García S, Rabinovich NR. Ivermectin and Novel Coronavirus Disease (COVID-19): Keeping Rigor in Times of Urgency.Am J Trop Med Hyg.2020; 102(6):1156–1157. DOI: 10.4269/ajtmh.20-0271.

6. Caly L, Druce JD, Catton MG, Jans DA, Wagstaff KM. The FDA-approved drug ivermectin inhibits the replication of SARS-CoV-2 in vitro. Antiviral Res. 2020; 178: 104787. DOI: 10.1016/j.antiviral.2020.104787.

7. European Medicines Agency. EMA advises against use of ivermectin for the prevention or treatment of COVID-19 outside randomised clinical trials. Published 22 March 2021. Accessed 23 March 2021. https://www.ema.europa.eu/en/news/ema-advises-against-use-ivermectin-prevention-treatment-covid-19-outside-randomised-clinical-trials

8. U.S. Food and Drug Administration. Why you should not use ivermectin to Treat or Prevent COVID-19? Published 3 May 2021. Accessed 5 May 2021. https://www.fda.gov/consumers/consumer-updates/why-you-should-not-use-ivermectin-treat-or-prevent-covid-19.

9. World Health Organization. Therapeutics and COVID-19: living guideline. WHO reference number: WHO/2019-nCoV/therapeutics/2021.

10. Bhimraj A, Morgan RL, Shumaker AH, Lavergne V, Baden L, Cheng VC, et al. Infectious Diseases Society of America Guidelines on the Treatment and Management of Patients with COVID-19. Infectious Diseases Society of America 2021; Version 4.2.0. Published 14 April 2021. Accessed 15 April 2021. https://www.idsociety.org/practice-guideline/covid-19-guideline-treatment-and-management/.

11. Padhy BM, Mohanty RR, Das S, Meher BR. Therapeutic potential of ivermectin as add on treatment in COVID 19: A systematic review and meta-analysis. J Pharm Pharm Sci. 2020;23:462–469. DOI: 10.18433/jpps31457.

12. Siemieniuk RA, Bartoszko JJ, Ge L, et al. Drug treatments for covid-19: living systematic review and network meta-analysis. BMJ. 2020; 370: m2980. DOI: 10.1136/bmj.m2980.

13. Kow CS, Merchant HA, Mustafa ZU, Hasan SS. The association between the use of ivermectin and mortality in patients with COVID-19: a meta-analysis. Pharmacol Rep. 29 Mar 2021;1–7. DOI: 10.1007/s43440-021-00245-z [Epud ahead of print].

14. Castañeda-Sabogal A, Chambergo-Michilot D, Toro-Huamanchumo CJ, et al. Outcomes of Ivermectin in the treatment of COVID-19: a systematic review and meta-analysis. 27 January 2021. medRxiv preprint. DOI: 10.1101/2021.01.26.21250420.

15. Hill A; International Ivermectin Project Team. Preliminary meta-analysis of randomized trials of ivermectin to treat SARSCoV-2 infection. medRxiv preprint. 19 Jan 2021. DOI: 10.21203/rs.3.rs-148845/v1.

16. Bryant A, Lawrie TA, Dowswell T, et al. Ivermectin for Prevention and Treatment of COVID-19 Infection: a Systematic Review and Meta-analysis. ResearchSquare preprint. 18 March 2021.DOI: https://doi.org/10.21203/rs.3.rs-317485/v1

17. Ivermectin for COVID-19: real-time meta-analysis of 54 studies. https://ivmmeta.com.Published8 May 2021. Accessed 9 May 2021.

18. Lawrie T. Ivermectin reduces the risk of death from COVID-19 -a rapid review and meta-analysis in support of the recommendation of the Front Line COVID-19 Critical Care Alliance. Published5 January 2021. Accessed 8 May 2021. DOI: 10.13140/RG.2.2.27751.88486.

19. Kory P, Meduri GU, Iglesias J, et al. Review of the Emerging Evidence Demonstrating the Efficacy of Ivermectin in the Prophylaxis and Treatment of COVID-19. FLCCC Alliance. Published 16 January 2021. Accessed 8 May 2021.https://covid19criticalcare.com.

20. WHO Working Group on the Clinical Characterization and Management of COVID-19 infection. A minimal common outcome measure set for COVID-19 clinical research. Lancet Infect Dis. 2020;20(8):e192–e197. doi:10.1016/S1473-3099(20)30483-7.

21. Sterne JA, Savović J, Page MJ, et al. RoB 2: a revised tool for assessing risk of bias in randomised trials. BMJ. 2019;366:4898.

22. Moher D, Liberati A, Tetzlaff J, Altman DG; PRISMA Group. Preferred reporting items for systematic reviews and meta-analyses: the PRISMA statement. PLoS Med. 2009;6(7):e1000097.

23. Veroniki AA, Jackson D, Viechtbauer W, et al. Methods to Estimate the between□Study Variance and Its Uncertainty in Meta□Analysis.Res Synth Methods. 2015;7(1):55–79.

24. Hartung J, Knapp G. A refined method for the meta-analysis of controlled clinical trials with binary outcome. Stat Med. 2001;20(24):3875–3889.

25. Sweeting MJ, Sutton AJ, Lambert PC. What to add to nothing? Use and avoidance of continuity corrections in meta-analysis of sparse data. Stat Med. 2004; 23: 1351–1375. DOI: 10.1002/sim.1761.

26. Balshem H, Helfand M, Schünemann HJ, et al. GRADE guidelines: 3. Rating the quality of evidence. J Clin Epidemiol. 2011;64(4):401–406. DOI: 10.1016/j.jclinepi.2010.07.015.

27. GRADEpro GDT: GRADEproGuideline Development Tool [Software]. McMaster University, 2020 (developed by Evidence Prime, Inc.). Accessed 7 May 2021. www.gradepro.org.

28. Chachar AZK, Khan KA, Asif M, Tanveer K, Khaqan A, Basri R. Effectiveness of Ivermectin in SARS-CoV-2/COVID-19 Patients. Int J Sci. 2020;9(09):31–35. DOI: 10.18483/ijSci.2378

29. Krolewiecki A, Lifschitz A, Moragas M, et al. Antiviral Effect of High-Dose Ivermectin in Adults with COVID-19: A Pilot Randomized, Controlled, Open Label, Multicentre Trial. SSRN; Jan 2020. DOI:10.2139/ssrn.3714649

30. Niaee MS, Gheibi N, Namdar P, et al. Ivermectin as an adjunct treatment for hospitalized adult COVID-19 patients: A randomized multi-center clinical trial. ResearchSquare.24 Nov 2020. DOI:10.21203/rs.3.rs-109670/v1.

31. Podder C, Chowdhury N, Sina M, Haque W. Outcome of ivermectin treated mild to moderate COVID-19 cases: a single-centre, open-label, randomized controlled study. IMC J Med Sci. 2020;14. doi:10.3329/imcjms.v14i2.52826.

32. Ahmed S, Karim MM, Ross AG, et al. A five-day course of ivermectin for the treatment of COVID-19 may reduce the duration of illness. Int J Infect Dis. 2021;103:214–216. doi:10.1016/j.ijid.2020.11.191.

33. Beltrán-Gonzalez JL, Gámez MG, Enciso EAM, et al. Efficacy and safety of Ivermectin and Hydroxychloroquine in patients with severe COVID-19. A randomized controlled trial. medRxiv. 23 Feb 2021. DOI:10.1101/2021.02.18.21252037

34. Chaccour C, Casellas A, Matteo AB-D, et al. The effect of early treatment with ivermectin on viral load, symptoms and humoral response in patients with non-severe COVID-19: A pilot, double-blind, placebo-controlled, randomized clinical trial. EClinicalMedicine. 2021;32: 100720. DOI:10.1016/j.eclinm.2020.100720.

35. Karamat HSB, Asma A, Najma P, et al. Efficacy of Ivermectin in COVID-19 Patients with Mild to Moderate Disease. medRxiv. 5 Feb 2021. DOI:10.1101/2021.02.02.21250840

36. López-Medina E, López P, Hurtado IC, et al. Effect of Ivermectin on Time to Resolution of Symptoms Among Adults With Mild COVID-19: A Randomized Clinical Trial. JAMA. 2021;325(14):1426–1435. doi:10.1001/jama.2021.3071.

37. Ravikirti, Roy R, Pattadar C, et al. Ivermectin as a potential treatment for mild to moderate COVID-19 – A double blind randomized placebo-controlled trial. medRxiv. 9 Jan 2021. DOI:10.1101/2021.01.05.21249310

38. Macdonald H, Loder E, Abbasi K. Living systematic reviews at The BMJ. BMJ. 2020;370:m2925. DOI: 10.1136/bmj.m2925.

39. Garegnani LI, Madrid E, Meza N. Misleading clinical evidence and systematic reviews on ivermectin for COVID-19. BMJ Evidence Based Medicine. 2021;1–3. DOI: 10.1136/bmjebm-2021-111678 [Epud ahead of print].

40. Forman R, Atun R, McKee M, Mossialos E. 12 Lessons learned from the management of the coronavirus pandemic. Health Policy 2020;124:577–580. DOI: 10.1016/j.healthpoi.2020.05.008.

41. Al Saidi Amo, Nur FA, Al-Mandhari AS, El Rabbat M, Hafeez A, Abubakar A. Decisive leadership is a necessity in the COVID-19 response. Lancet 2020;396(10247):295–298. DOI: 10.1016/S0140-6736(20)31493-8.

42. Scheufele DA, Hoffman AJ, Neeley L, Reid CM. Misinformation about science in the public sphere. PNAS 2021;118(15):e2104068118. DOI: 10.1073/pnas.2104068118.

43. Walker E, Hernandez AV, Kattan MW. Meta-analysis: its strengths and limitations. Clev Clin J Med. 2008;75(6):431–439.

44. Hernandez AV, Marti KM, Roman YM. Meta-analysis. Chest 2020;158(1S):S97–S102. DOI: https://doi.org/10.1016/j.chest.2020.03.003.

