## Supplemental File for "Ivermectin for the treatment of COVID-19: A systematic review and meta-analysis of randomized controlled trials"

Yuani M. Roman et al.

**Pubmed search strategy**

**Figure S1**. Cochrane Risk of Bias 2.0 Tool of included randomized controlled trials.

**Figure S2.** Subgroup analyses

Fig S2.1. All-cause mortality

S2.1.a. Severity

S2.1.b. RoB

Fig S2.2. Length of stay

S2.2.a. Severity

S2.2.b. RoB

Fig S2.3. Adverse events

S2.3.a. Severity

S2.3.b. RoB

Fig S2.4. Severe adverse events

S2.4.a. Severity

S2.4.b. RoB

Fig S2.5. Viral clearance

S2.5.a. Severity

S2.5.b. RoB

**Figure S3.** Sensitivity analyses

Fig S3.1. All-cause mortality

Fig S3.2. Length of stay

**Table S1.** Systematic reviews evaluating the efficacy of ivermectin for the treatment of COVID-19that were published as full text.

**Table S2.** Systematic reviews and meta-analyses evaluating the efficacy of ivermectin for the treatment of COVID-19, and published only as pre-prints and online resources.

**PubMed search strategy**

("ivermectin"[MeSH Terms] OR "ivermectin"[All Fields] OR "ivermectine"[All Fields] OR "ivermectin s"[All Fields] OR "ivermectins"[All Fields]) AND ("covid 19"[All Fields] OR "covid 19"[MeSH Terms] OR "covid 19 vaccines"[All Fields] OR "covid 19 vaccines"[MeSH Terms] OR "covid 19 serotherapy"[All Fields] OR "covid 19 serotherapy"[Supplementary Concept] OR "covid 19 nucleic acid testing"[All Fields] OR "covid 19 nucleic acid testing"[MeSH Terms] OR "covid 19 serological testing"[All Fields] OR "covid 19 serological testing"[MeSH Terms] OR "covid 19 testing"[All Fields] OR "covid 19 testing"[MeSH Terms] OR "sarscov 2"[All Fields] OR "sarscov 2"[MeSH Terms] OR "severe acute respiratory syndrome coronavirus 2"[All Fields] OR "ncov"[All Fields] OR "2019 ncov"[All Fields] OR (("coronavirus"[MeSH Terms] OR "coronavirus"[All Fields] OR "cov"[All Fields])

**Figure S1. Cochrane Risk of Bias 2.0 Tool of included randomized controlled trials**

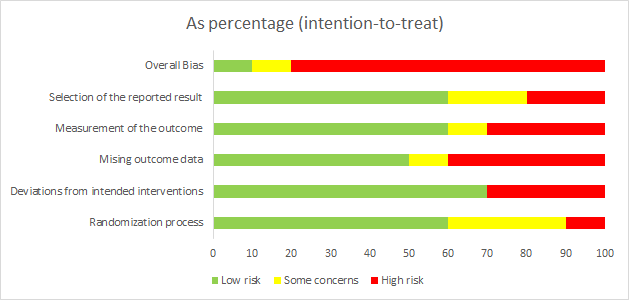

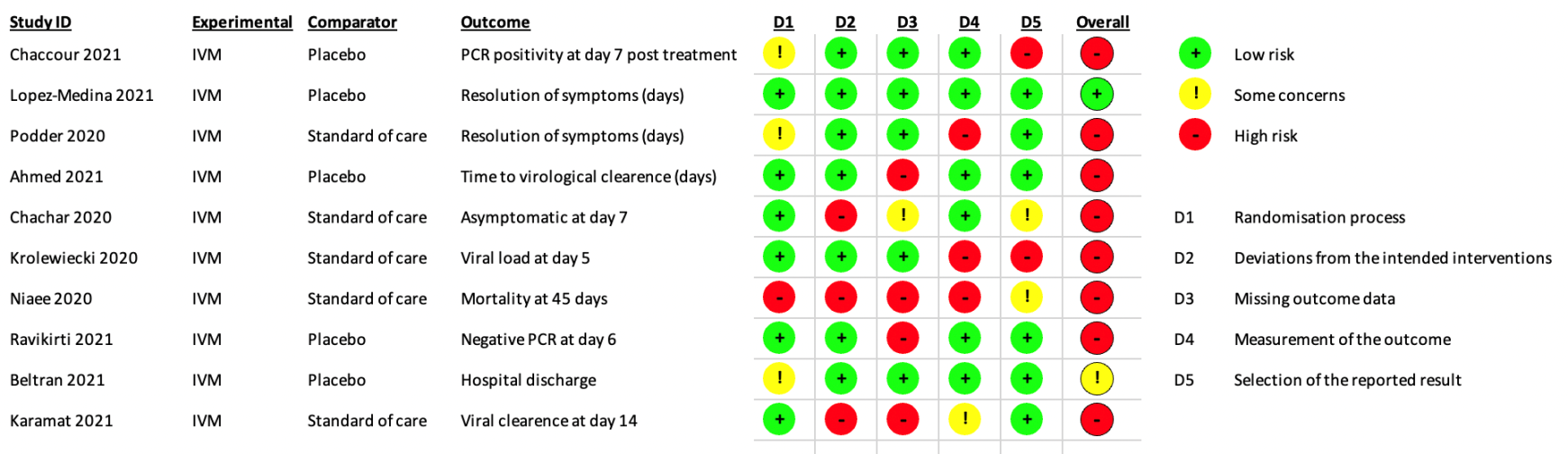

**Figure S2. Subgroup analyses**

**Fig S2.1. All-cause mortality**

**S2.1.a. Severity**

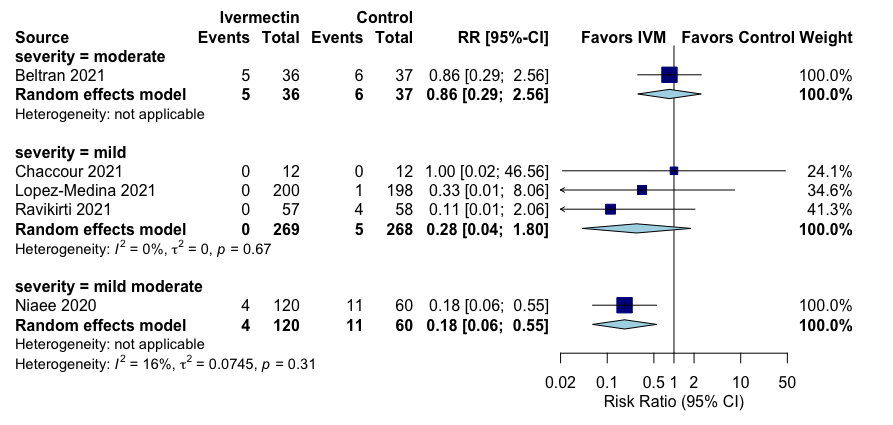

**S2.1.b. RoB**

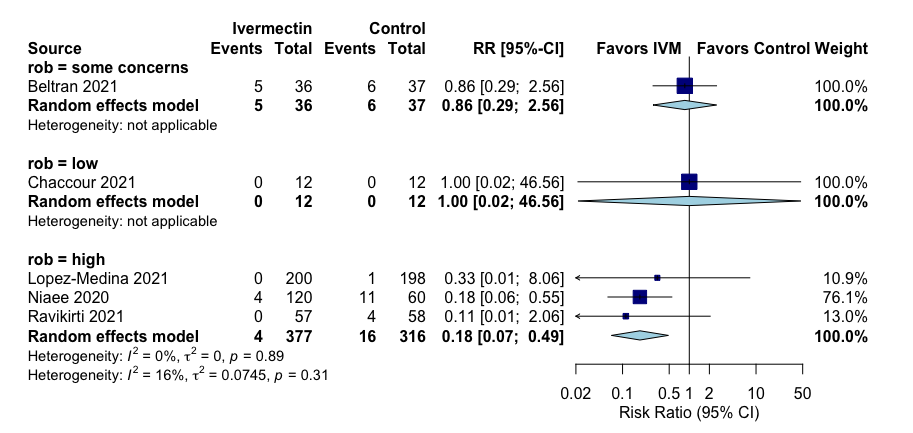

**Fig S2.2. Length of stay**

**S2.2.a. Severity**

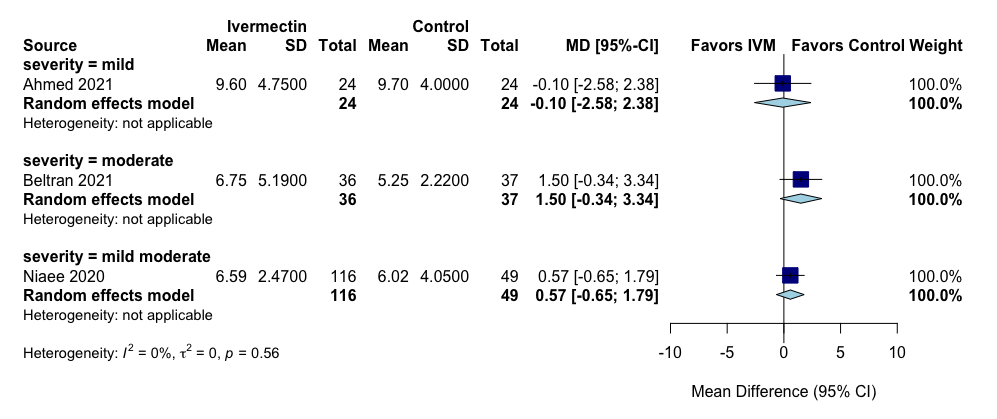

**S2.2.b. RoB**

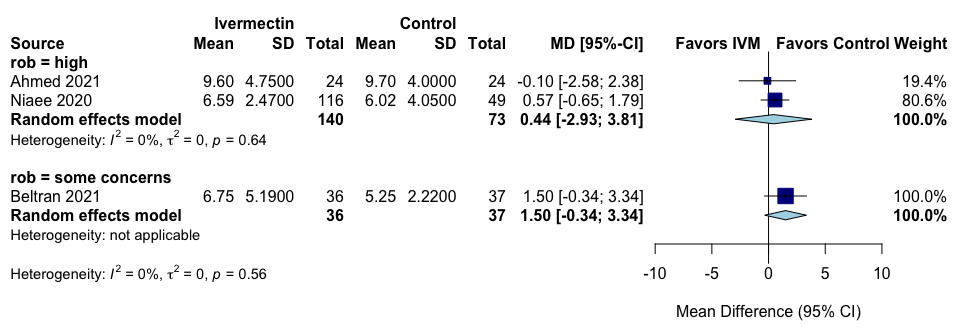

**Fig S2.3. Adverse events**

**S2.3.a. Severity**

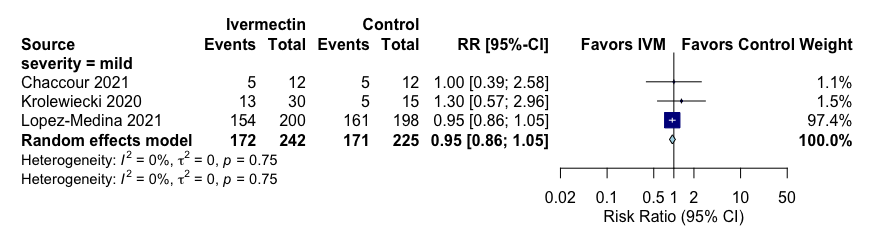

**S2.3.b. RoB**

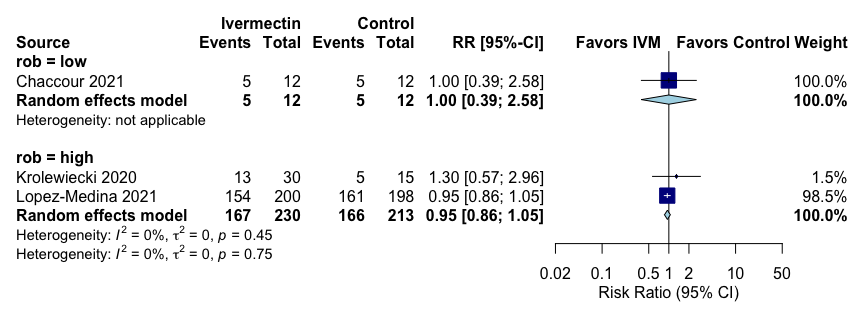

**Fig S2.4. Severe adverse events**

**S2.4.a. Severity**

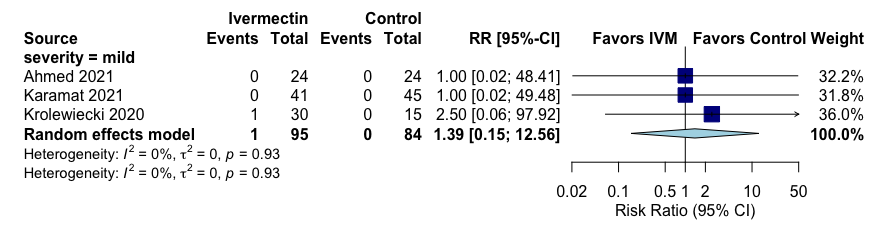

**S2.4.b. RoB**

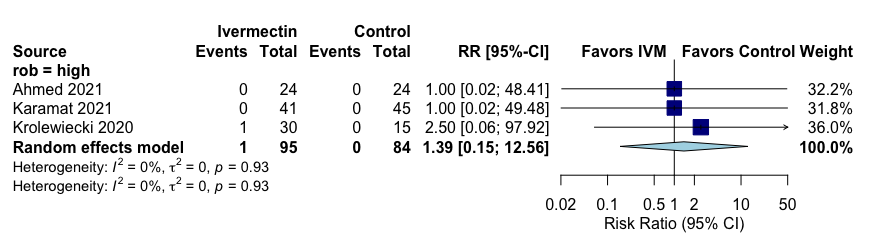

**Fig S2.5. Viral clearance**

**S2.5.a. Severity**

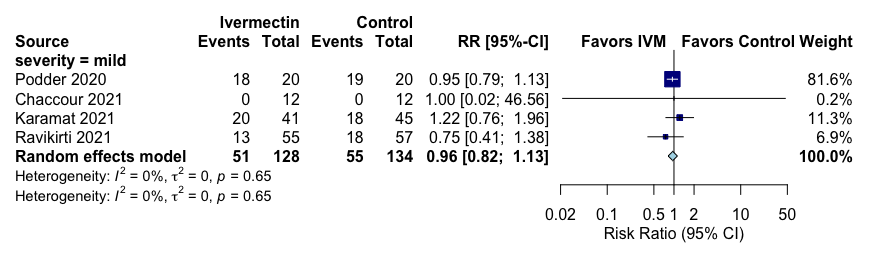

**S2.5.b. RoB**

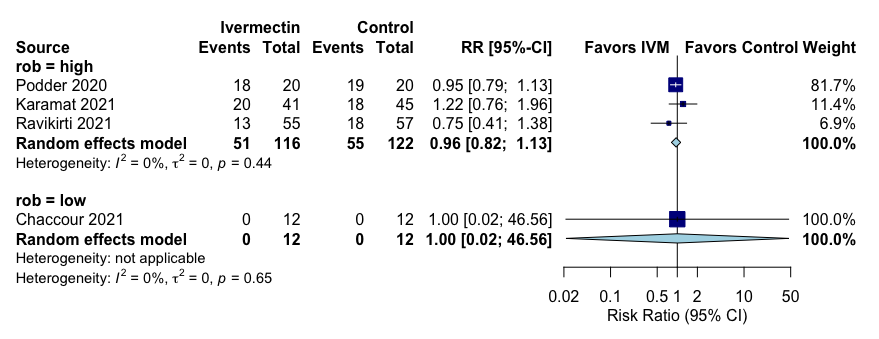

**Figure S3. Sensitivity analyses**

**Fig S3.1. All-cause mortality**

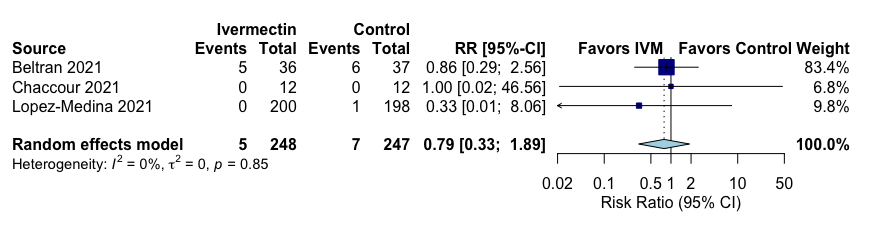

**Fig S3.2. Length of stay**

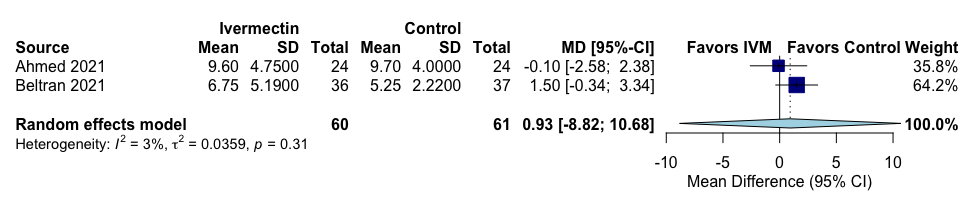

**Table S1.** Systematic reviews evaluating the efficacy of ivermectin for the treatment of COVID-19that were published as full text.

| **Characteristics** | **Siemieniuk et al., 2021^12^** | **Padhy et al., 2020^11^** | **WHO, 2021^9^** | **Kow et al., 2021^13^** | **Our study** |
| --- | --- | --- | --- | --- | --- |
| Type of review | Living systematic review with network meta-analysis. | Systematic review and meta-analysis. | Living systematic review and network meta-analysis | Systematic review and meta-analysis | Systematic review with meta-analysis. |
| Type of publication | Full text | Full text | Full text | Full text | Current |
| Primary objectives | To compare the effects of treatment for COVID-19 | To assess the currently available data on the therapeutic potential of ivermectin for the treatment of COVID‐19 as add on therapy. | To evaluate the role of drugs in the treatment of patients with COVID-19 | To investigate the mortality benefits of ivermectin in patients with COVID-19. | To evaluate the effects of ivermectin as an antiviral treatment on death and recovery in people with COVID-19. |
| Primary outcome(s) | Mortality; mechanical ventilation; admission to hospital; adverse effects leading to discontinuation; viral clearance; duration of hospital stay; ICU length of stay; duration of mechanical ventilation; time to symptom resolution; time to viral clearance; ventilator free days. | All-cause mortality and any death during the available period of follow up in the studies. | Outcomes [rating from 1 (not important) to 9 (critical)] taking a patient's perspective. The panel prioritized outcomes from both an inpatient and outpatient perspective. | All-cause mortality. | All-cause mortality, length of hospital stay, adverse events, severe adverse events, viral clearance. |
| Inclusion criteria | Randomized clinical trials in people with suspected, probable, or confirmed COVID-19 that compared drugs for treatment against one another or against no intervention, placebo, or standard care. | Randomized controlled trials and observational studies. | Randomized controlled trials. | Randomized controlled trials. | Randomized controlled trials. |
| Databases searched | WHO COVID-19 database, CDC COVID-19 Research Articles Downloadable Database, MEDLINE (Ovid and Pubmed), PubMed Central, Embase, CAB Abstracts, Global Health, PsycInfo, Cochrane Library, Scopus, Academic Search Complete, Africa Wide Information, CINAHL, ProQuest Central, SciFinder, the Virtual Health Library, LitCovid, WHO covid-19 website, CDC covid-19 website, Eurosurveillance, China CDC Weekly, Homeland Security Digital Library, ClinicalTrials.gov, bioRxiv (preprints), medRxiv (preprints), chemRxiv (preprints), and SSRN (preprints). | PubMed, EMBASE, the Cochrane Library, SCOPUS, Web of Science, the reference lists of retrieved articles were checked for additional studies. For unpublished data, International Clinical Trials Registry Platform (ICTRP), ClinicalTrials.gov and pre-print servers medRxiv and bioRxiv were also searched. | Not described. | PubMed, Google Scholar, Cochrane Central Register of Controlled Trials, and preprint servers (medRxiv, Research Square, SSRN). | OVID-MEDLINE, OVID-EMBASE, Scopus, and Cochrane Library. |
| Number of studies included | Mortality: 7 trials; mechanical ventilation: 4 trials; adverse effects leading to discontinuation: 3 trials; viral clearance at 7 days: 7 trials; length of hospital stay: 3 trials; time to resolution of symptoms: 2 trials; time to viral clearance: 3 trials; admission to hospital: 1trial. | 4 observational studies (3 with control arm and 1 without control arm). The pooled effect of add on ivermectin was evaluated in the 3 controlled studies. | 16 randomized controlled trials | 6 randomized controlled trials | 10 randomized controlled trials. |
| Sample size | Mortality: 751 participants; mechanical ventilation: 407 participants; adverse effects leading to discontinuation: 270 participants; viral clearance at 7days: 291 participants; length of hospital stay: 178 participants; time to resolution of symptoms: 260 participants; time to viral clearance: 234 participants; admission to hospital: 200 participants. | 629 participants.  481 participants in the 3 controlled studies. | 2407 participants | 1255 participants | 1173 participants |
| Quality assessment (risk of bias in individual studies) | Cochrane tool for randomized trials (RoB 2.0). | ROBINS-I for non-randomized studies of interventions. | Not described. | Cochrane tool for randomized trials (RoB 2.0). | Cochrane tool for randomized trials (RoB 2.0). |
| Methods of meta-analysis | Random-effects models. | Random-effects models. | Random-effects models. | Random-effects models and inverse variance heterogeneity model for the meta-analysis to estimate the pooled odds ratio at 95% confidence intervals. | Random-effects models. |
| Definition of heterogeneity | Not specified. | Heterogeneity was evaluated by I^2^. | Not described. | Heterogeneity was evaluated by I^2^. | Heterogeneity was evaluated by I^2^. |
| Outcome effects with 95% CI | Risk difference per 1000 vs. standard of care:  -Mortality: -103 (95%CI: -117 to -78) -Mechanical ventilation: -54 (95%CI: -100 to 80) -Admission to hospital: -32 (95%CI: -47 to 23) -Adverse effects leading to discontinuation: 26 (95%CI -2 to 187) -Viral clearance: 118 (95%CI -13 to 241) -Duration of hospital stay: -0.5 (95%CI -1.7 to 1.1) -Time to symptom resolution: -0.4 (95%CI -3.7 to 3.5) -Time to viral clearance: -2.0 (-4.4 to 2.4) | -All-cause mortality: OR 0.53 (95%CI: 0.29 to 0.96) | -Mortality: OR 0.19 (95%CI 0 .09 - 0.36) | -Mortality: OR 0.21(95%CI 0.11 - 0.42). | -All-cause mortality:  RR 1.11 (95%CI 0.16 to 7.65)  -Length of stay:  MD 0.72 days, 95%CI -0.86 to 2.29  -Adverse events:  RR 0.95, 95%CI 0.85 to 1.07  -Severe adverse events:  RR 1.39, 95%CI 0.36-5.30  -Viral clearance:  RR 0.96, 95CI 0.79-1.16 |

**Table S2.** Systematic reviews and meta-analyses evaluating the efficacy of ivermectin for the treatment of COVID-19, and published only as pre-prints and online resources.

| **Characteristics** | **Castañeda-Sabogalet al., 2021^14^** | **Hill et al., 2021^15^** | **Bryant et al., 2021^16^** | **https://ivmmeta.com/ 2021^17^** | **Kory et al., 2021^19^** | **Lawrie et al., 2021^18^** |
| --- | --- | --- | --- | --- | --- | --- |
| Type of review | Systematic review with meta-analysis. | Systematic review and meta-analysis. | Systematic review with meta-analysis. | “Real time meta-analysis”. | Meta-analysis. | “Rapid systematic review and meta-analysis”. |
| Type of publication | Pre-print. | Pre-print. | Pre-print. | On-line. | On-line. | On-line. |
| Primary objectives | To assess the outcomes of ivermectin in ambulatory and hospitalized patients with COVID-19. | To combine available results from published or unpublished randomized trials of ivermectin in SARS-CoV-2 infection. | To assess the efficacy of ivermectin treatment among people with COVID-19 infection. | Not available. | Not available. | Not described. |
| Primary outcome | Overall mortality. | All-cause mortality. | Death from any cause. | Not available but more serious outcomes had priority. | Time to clinical recovery and mortality. | Death. |
| Inclusion criteria | All original published studies (either as preprints or in scientific journals) of clinical trials, non-randomized studies of intervention, and retrospective cohorts. | Randomized clinical trials. | Randomized clinical trials (double blind and open label) and quasi-randomized clinical trials. | All studies regarding the use of ivermectin for COVID-19 that report an effect compared to a control group. | Not available. | Randomized clinical trials and controlled observational studies. |
| Databases searched | PubMed, Scopus, Web of Science, Ovid-Medline, Embase, websites for preprints/preproofs (“Other sources”; https://www.medrxiv.org, https://preprints.scielo.org/index.php/scielo, https://www.biorxiv.or, https://arxiv.org), websites for protocols of clinical trials (https://clinicaltrials.gov). | PUBMED, EMBASE, MedRxiv, clinicaltrials.gov, WHO International Clinical Trials Registry Platform (ICTRP), and Stanford University’s Coronavirus Antiviral Research Database (CoV-RDB), to identify additional trials listed on other national, and international registries. | Medline, Embase, CENTRAL, Cochrane COVID-19 Study Register and Chinese databases; the reference list of included studies and of two other 2021 literature reviews on ivermectin; contact with experts in the field; all trials registered on clinical trial registries were checked and trial ists of 39 ongoing trials or unclassified studies were contacted to request information; Medrix and the International Clinical Trials Registry Platform. | PubMed, medRxiv, ClinicalTrials.gov, The Cochrane Library, Google Scholar, Collacovid, Research Square, ScienceDirect, Oxford University Press, the reference lists of other studies and meta-analyses, and submissions to the site c19ivermectin.com, | Not available. | Not available. Summary tables of the Front Line COVID-19 Critical Care Alliance (FLCCC)  (https://covid19criticalcare.com) |
| Number of studies included | 12 studies (6 randomized controlled trials; 5 cohorts, 1 case series) (5 were published studies). | 18 randomized controlled trials (5 were published studies). 6 studies evaluated survival. | 19 randomized controlled trials contributed data to the comparison ivermectin treatment vs no ivermectin treatment for COVID-19 treatment.  13 randomized controlled trials included in the meta-analysis. | 46 studies, including 24 randomized controlled trials.  16 studies (10 randomized controlled trials) to early treatment.  19 studies (11 randomized controlled trials) to late treatment. | 8 randomized controlled trials evaluated time to clinical recovery.  10 studies (4 randomized controlled trials and 6 observational studies) evaluated mortality. | 12 studies, 9 randomized controlled trials and 3 controlled observational studies. |
| Sample size | 7412 participants. | 2282 participants.  1255 participants in the 6 studies evaluating survival. | 2003 participants.  1892 participants included in the meta-analysis. | 15480 participants.  Early treatment: 1684. participants.  Late treatment: 6785. participants. | 1054 participants to evaluate the time to clinical recovery.  3478 participants to evaluate mortality. | 1835 participants. |
| Quality assessment (risk of bias in individual studies) | ROBINS-I for non-randomized studies of interventions and the Cochrane tool for assessing risk of bias in randomized trials (RoB 2.0). | Cochrane tool for assessing risk of bias in randomized trials. | Cochrane tool for assessing risk of bias in randomized trials. | Not available.  The authors cited that in order to avoid bias in the selection of studies, they included all studies in the main analysis. | Not available. | ROBINS-I for non-randomized studies of interventions and the Cochrane handbook for randomized trials. |
| Methods of meta-analysis | Random-effects models and the inverse variance method. | Random-effects and the inverse-variance method. | Random-effects models and the inverse variance method. | Random-effects models. | Not available. | Random effects models. |
| Definition of heterogeneity | Heterogeneity was evaluated by I^2^. | Heterogeneity was evaluated by I^2^. | Heterogeneity was evaluated by I^2^. | Not available | Not available. | Heterogeneity was evaluated by I^2^ and was assessed by visual inspection of forest plots. |
| Outcome effects with 95% CI | - Mortality: RR 0.70 (95%CI 0.31 to 2.28)  - Recovery:  RR 1.37 (95%CI 0.61 to 3.07). | -All-cause mortality: RR 0.25 (95%CI 0.12 to 0.52). | -Death for any cause: Reduced the risk of death by an average of 68% (95% CI, 28–86%).  aRR 0.32 (95% CI 0.14 to 0.72). | -Mortality: RR 0.25 (95%CI 0.15 to 0.44) for all treatment delays.  RR 0.16 (95%CI 0.04 to 0.63) for early treatment.  - Improvement: RR 0.30 (95%CI 0.19 to 0.47). | -Mortality: OR 0.29 (95%CI 0.19 to 0.45). | -Mortality: RR 0.17 (95% 0.08 to 0.35). |
